# Modeling the Transmission of Respiratory Infectious Diseases in Mass Transportation Systems

**DOI:** 10.1101/2020.06.09.20126334

**Authors:** Christian Alvin H. Buhat, Destiny SM. Lutero, Yancee H. Olave, Monica C. Torres, Jomar F. Rabajante

## Abstract

Mass transportation is one of the areas that are badly hit by respiratory infectious disease outbreaks due to moderate to high exposure risk to pathogens brought about by the interaction among commuters. Here, we formulate agent-based models that simulate the spread of a respiratory infectious disease in a train wagon in the Manila Light Rail Transit System, and in a 49-seater public utility bus. We consider preventive measures such as implementation of social distancing, and limitation of interaction or movement among the commuters to investigate how these measures will inhibit disease transmission. We also consider the effect of protective gears and practices, crowd density, and prevalence of disease in the community on the possible number of newly-infected individuals. Our simulations show that (i) individuals must have protection with more than 90% effectiveness to inhibit transmission of the disease; (ii) social or physical distancing by more than 1m distance reduces the risk of being infected; (iii) minimizing movement or interaction with other passengers reduces the risk of transmission by 50%; (iv) passenger capacity should be less than 10-50% of the maximum seating capacity to reduce the number of infections depending on the level of imposed social distancing and passenger interaction; (v) vehicles with greater number of occupied seating capacity generate higher number of infections but vehicles with smaller dimensions have faster disease transmissions; and (vi) ideal set-up for a 24-seater train wagon (49-seater bus) is to allow a maximum of 12 (24) passengers, with little to no interaction among passengers, with social distancing of more than 1m distance apart, and each person has a protection with 90% effectiveness as much as possible. These results can aid policy makers in determining optimal strategies to minimize infections while maintaining transportation services during pandemics or disease outbreaks.

## 1. Introduction

Several respiratory diseases have spread and caused deaths globally. The 1918 influenza that resulted in at least 50 million deaths worldwide was the most severe pandemic in recent history [7]. During the pandemic, some non-pharmaceutical interventions including school closures, prohibition of public gatherings, and isolation or quarantine orders were employed [7]. This pandemic was followed by two other worldwide influenza outbreaks in 1957 and 1968 [9]. In 2003, severe acute respiratory syndrome (SARS) which is caused by a member of the betacoronavirus subgroup named SARS-CoV had spread rapidly worldwide and resulted in 8422 cases with 916 deaths [3, 15]. Control measures that were imposed for SARS include case detection and isolation, contact tracing, quarantine, physical distancing, increased personal hygiene, wearing of mask, and screening travelers at borders [3, 2]. Recently, there has been a worldwide outbreak of another member of the beta group of coronaviruses named SARS-CoV-2 [15]. The disease is called coronavirus disease 19 (COVID-19). The aforementioned diseases are highly contagious and can be transmitted to other humans through exposure to coughing, sneezing, and respiratory droplets from an infected individual.

The World Health Organization (WHO) declared on March 11, 2020 that COVID-19 can be characterized as a pandemic [11]. Its worldwide spread in 2020 has affected public transportation systems of different countries. Growing concerns of infection due to interaction of infected individuals with the general population have driven changes in mobility and public transportation. Many countries, including countries in Asia such as China, India, Malaysia and Philippines, imposed restrictions or suspension on their public transportation to prevent the spread of the disease [16, 10, 14, 13]. In the Philippines, all kinds of public transportation including railway systems have been suspended in Luzon since March 17, 2020 in connection with the implementation of the enhanced community quarantine (ECQ) [8].

To slowly reopen economies, community lockdown restrictions are relaxed in several countries. Some establishments are allowed to operate which entails that some or all modes of public transportation are also allowed to function. In the presence of a pandemic, the public is still advised to be cautious, that is, strict physical distancing and other preventive measures must still be observed in workspaces and in public places including public transportation.

In the presence of such highly contagious respiratory diseases like COVID-19, while it is vital to sustain economical activities, infections must be maintained at a controllable level. Thus, combined effort of government and the public is needed to control transmission. Agent-based models (ABM) have been helpful to investigate dynamics of infectious diseases. Perez and Dragicevic proposed an agent-based model that incorporates geographic information systems (GIS) to simulate the outbreak of an infectious disease in an urban area [12]. The model was applied to study measles outbreak in an urban region [12]. Hackl and Dubernet linked a large-scale agent-based transport simulation with a generic epidemic spread model to simulate the spread of infectious diseases in an urban setting, and the model was used to examine seasonal influenza outbreak within the metropolitan area of Zurich [5]. A data-driven agent-based model was developed by Hunter et al. to simulate the spread of an airborne infectious disease, and was tested by simulating a 2012 measles outbreak in Schull, Ireland [6].

In this study, an agent-based model is developed to examine transmission of highly contagious respiratory diseases in mass transportation systems, specifically bus and train. Protection and crowd density levels are varied to determine the effect of preventive measures and limiting vehicle’s seating occupancy rate during the epidemic period. This paper is organized as follows. The *Model* gives information on how simulations work, buttons’ functions, and how simulations can be performed. The *Simulation Results* section discusses the impact of crowd density and protection against infection for both bus and train. Key messages are presented in the *Conclusion* part. The model can be further studied and improved by assessing the *Limitations* section.

## 2. Model Framework

We use an agent-based model (ABM) to simulate the transmission of a respiratory infectious disease in a mass transportation system. ABM is a micro-scale model used to demonstrate movements and interactions among agents in a complex system with the purpose of simulating their behavior [4]. It has been extensively used in various fields such as in biology to study population dynamics, and simulate the interaction among the individuals in the population [1].

Using the NetLogo programming environment for our ABM, we consider a 24-seater train wagon mimicking the situation in a segment of a train car in the Manila Light Rail Transit System, and a 49-seater public utility bus having 2 adjacent seats in each side per row for 11 rows and a row of 5 seats in the last. We also consider the coronavirus disease 19 (COVID-19), an infectious disease declared as pandemic in March 2020, as an inspiration. The model explores the effects of factors such as crowd density, protection level of individuals against infection, and initial number of infectious individuals. We determine these effects under the presence or absence of social distancing, and interaction among the passengers.

Initially, an individual is either infectious or non-infectious. An initially non-infectious individual will become infectious through increase in percentage of exposure to an infectious individual. In the model, individuals occupy vacant seats first and those left without seats will be standing. Their location in the vehicle will determine their percentage of exposure. A person within 2 meters of an infectious individual will have a 50% chance of being exposed while a person within 1 meter of an infectious individual will have 100% chance of being exposed. Exposed individuals may or may not become infected depending on their protection. Protection (hand washing, use of alcohols or sanitizers, wearing of face masks) influences the chance of an exposed individual to become unexposed, uninfected, or infected. If an exposed individual is not equipped with ample protection, he*/*she becomes infectious. Infectious individuals are infectious to other people and may transmit virus through the environment.

The **SETUP** button generates individuals according to the initial parameter values. Each individual has a chance of initially being infectious depending on the INIT-INF value. Once the model is set, the GO button starts the simulation and runs it continuously until: (1) all individuals are infectious, (2) no new infections can occur, or (3) maximum time step is reached. Each time step (tick) is considered as a 10-minute duration in the model.

Crowd density greater than 100% means that the number of passengers exceeds the set maximum. If social distancing is implemented: (i) for the train, passengers will be seated with 1 seat apart, and (ii) for the bus, passengers will be seated near the window side, and the adjacent seat will be vacant. The following table describes the variables of the model:

| Variable | Description | Value |
| --- | --- | --- |
| Maximum Capacity | Measure of maximum capacity of the vehicle | 24 for train, 49 for bus |
| Crowd Density | Density of individuals in the transportation system | 0 to 200% |
| Protection | Chance for an exposed individual to be back to being susceptible again, that is, unsuccessful transmission of the virus | 0 to 100% |
| Init-inf | Initial percentage of infectious individuals | 0 to 50% |
| Social Distancing? | Determines if individuals will try to stay with minimum of 1m apart and will observe social distancing protocol | Yes/No |
| Interact? | Determines if individuals will randomly interact with each other | Yes/No |

## 3. Simulation Results

We vary key parameters in the model and determine the effect of each parameter by looking at the resulting number of newly infected individuals. We observe the dynamics of transmission under four different scenarios (activation or non-activation of social distancing and interaction) for 100 minutes (10 ticks).

### 3.1. Protection against Infection

With a fixed crowd density of 50% on both mass transportation systems, we observe the effect of increasing the protection level of individuals to the number of new infected individuals.

#### 3.1.1. Inside a Train wagon

Figure 2 shows that minimizing interaction (due to random movement) among the passengers has a greater effect on reducing the number of new infected individuals. Minimizing the movement of passengers inside the train, coupled with social distancing, should be observed. Social distancing is not enough as the virus can be present in the environment.

**Figure 1:**
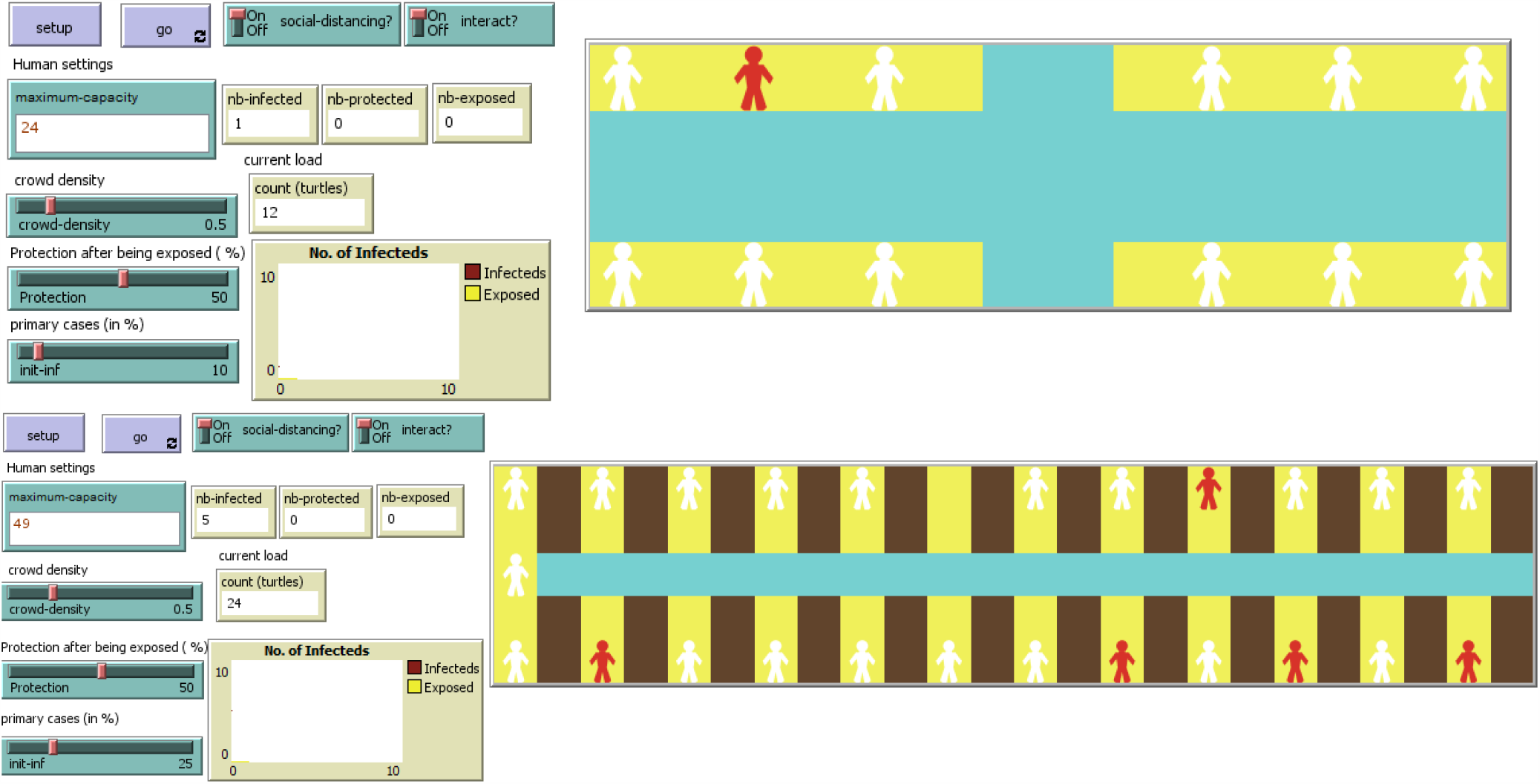
NetLogo environment of respiratory infectious disease transmission simulation inside a train wagon (top), and a bus (bottom)

**Figure 2:**
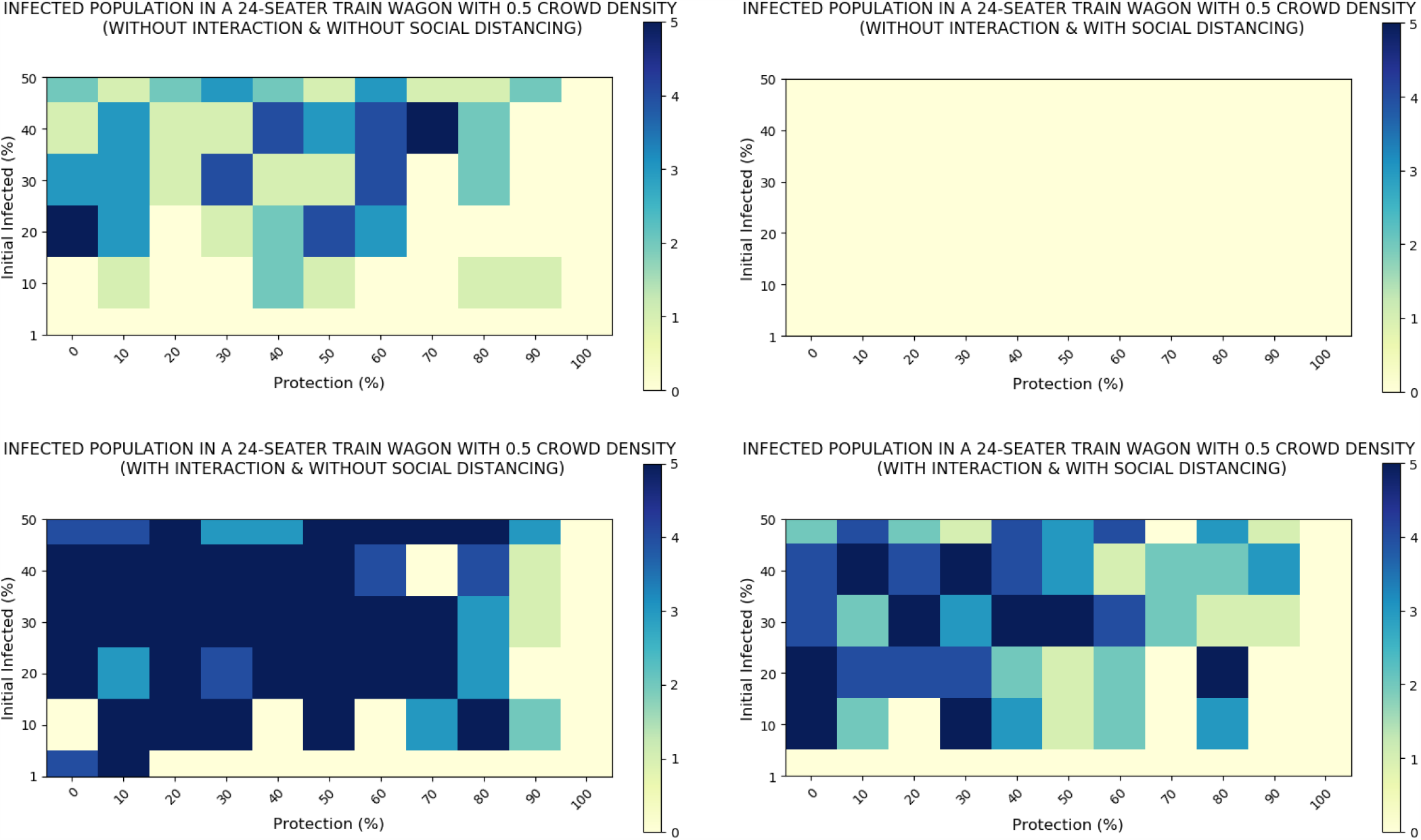
Inside a train: Number of new infected individuals based on the protection level (in %) and the initial percentage of infectious individuals (in %), with crowd density *=* 50%. The colors denote the number of new infected individuals.

The number of new infected individuals is least when social distancing is practiced and there is no interaction as represented by the lightest shade among the four heat maps in Figure 2. In fact, there is no new infected individual when crowd density is 50% or less under social distancing without interaction scenario. Likewise, number of new infected individuals is greatest when there is interaction and when social distancing is not practiced.

In all scenarios, given the same percentage of initial infected individuals, the number of new infected individuals decreases as protection increases. In a location where the disease is prevalent (e.g., possible number of initially infected individuals entering the train is likely more than one), high (90%) protection level is needed.

#### 3.1.2. Inside a Bus

Figure 3 shows that interaction appears to have a greater effect on the number of newly infected individuals compared to social distancing since the change in shade in the heat map (lighter to darker) is more significant when varying the interaction than just social distancing. More cases (greater than 5 newly infected) occur in cases with random passenger interaction.

**Figure 3:**
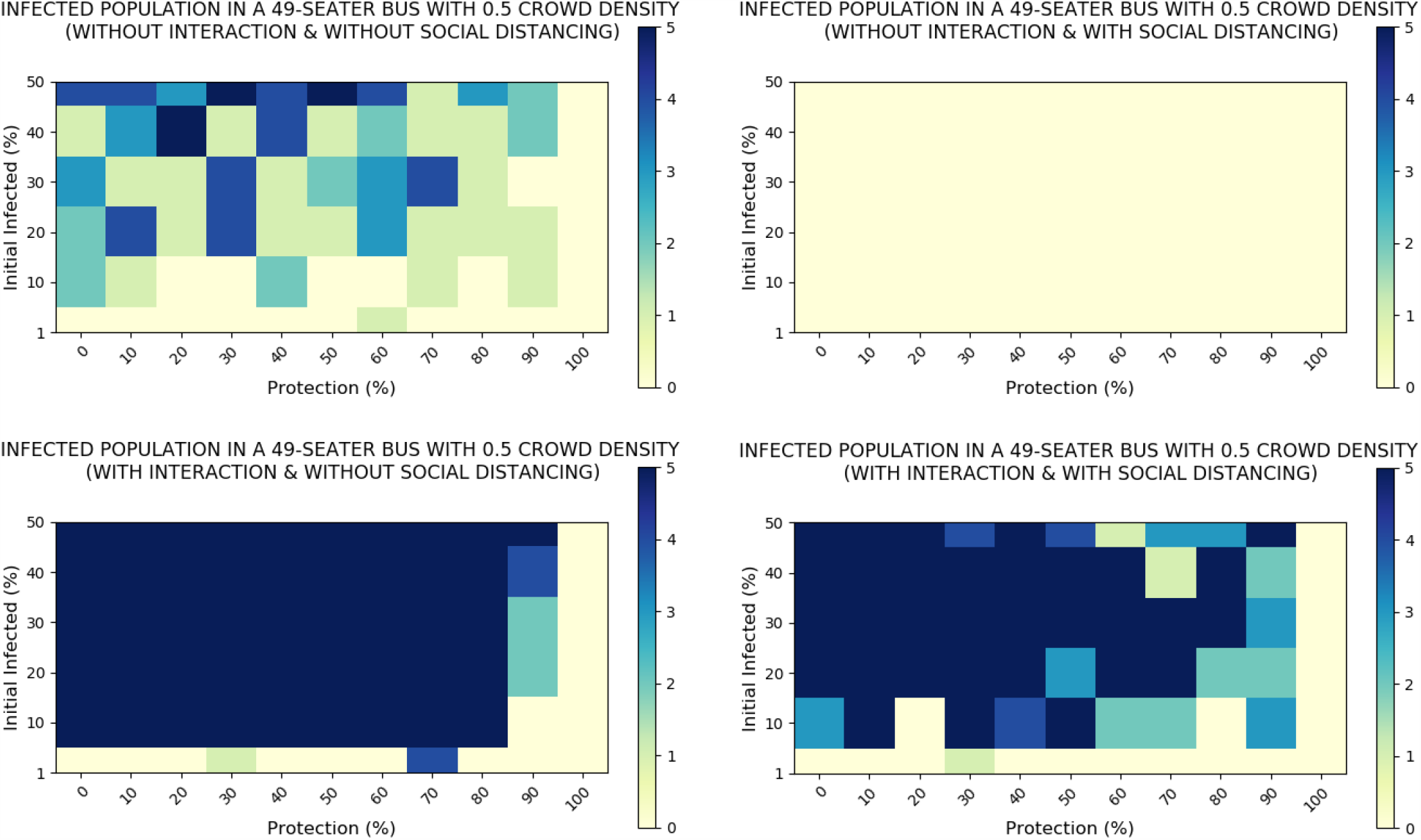
Inside a bus: Resulting final number of newly infected individuals based on the protection rate of individuals (in %) and the initial percentage of infected individuals (in %), with crowd density *=* 50%

Number of newly infected is least when there is no interaction and social distancing is practiced as represented by the lightest shade among the four heat maps in Figure 3. In fact, there are no newly infected individuals when crowd density is 0.5 or less. Likewise, number of newly infected is greatest when there is interaction and social distancing is not practiced. In all scenarios, given the same initial percentage of infected individuals, the number of new infected individuals decreases as protection increases.

### 3.2. Effect of Crowd Density

In the previous section, we observed that protection level affects the number of newly infected individuals. Now we look at the effect of crowd density inside a train wagon and a bus, observed with low (30%), and high (70%) protection rate. Note that in this section, protocol on social distancing is implemented (i.e., persons are more than 1m apart and one seat apart) but as soon as the number of passengers exceeds the maximum capacity, the model assumes that the extra passengers will stand in the aisle.

#### 3.2.1. Inside a Train wagon

Figure 4 shows that as the train wagon gets crowded (or overcrowded), the number of new infected individuals increases, as expected. To be conservative, crowd density should be less than 10% of the maximum capacity of the train wagon to ensure low risk of virus transmission. In some cases, e.g., when prevalence in the location is low, this proposed crowd density limit can be relaxed.

**Figure 4:**
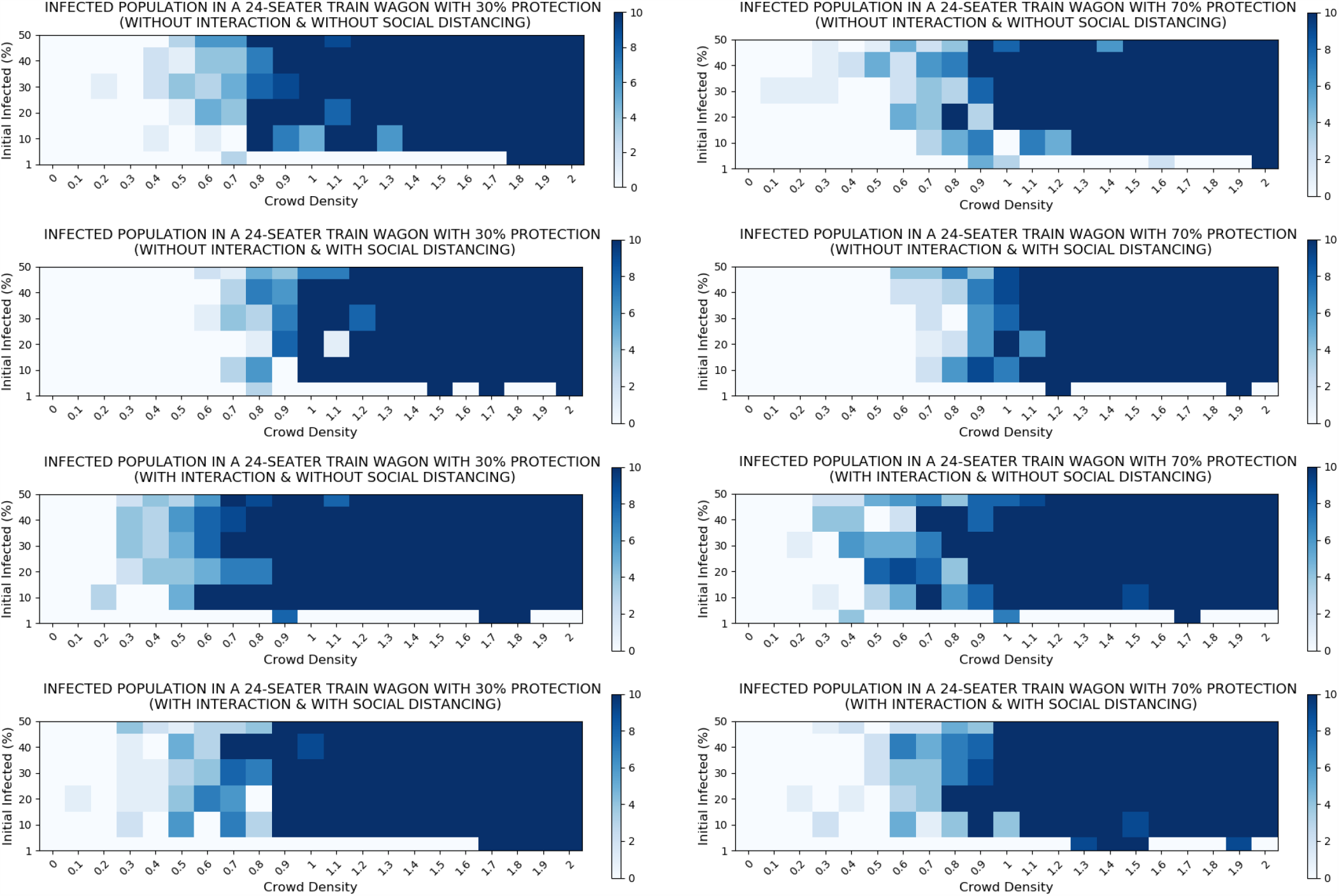
Inside a train: Resulting final number of newly infected individuals based on crowd density of passengers and the initial percentage of infected individuals (in %), with protection *=* 30% (left column), and *=* 70% (right column).

#### 3.3.2. Inside a Bus

Figure 5 shows that as the bus gets crowded (or overcrowded) the greater the number of individuals are being infected. Number of new infected is least up to 100% crowd density when social distancing is practiced and interaction is not allowed. Other than this case, crowd density should be less than 10% to prevent spread of infection. Likewise, number of newly infected is greatest when there is interaction and social distancing is not practiced. In general, as the crowd density increases, number of new infected increases. Also, overcrowding will result in a large number of infection in the whole vehicle as long as there is an infectious individual. These observations are true for all cases.

**Figure 5:**
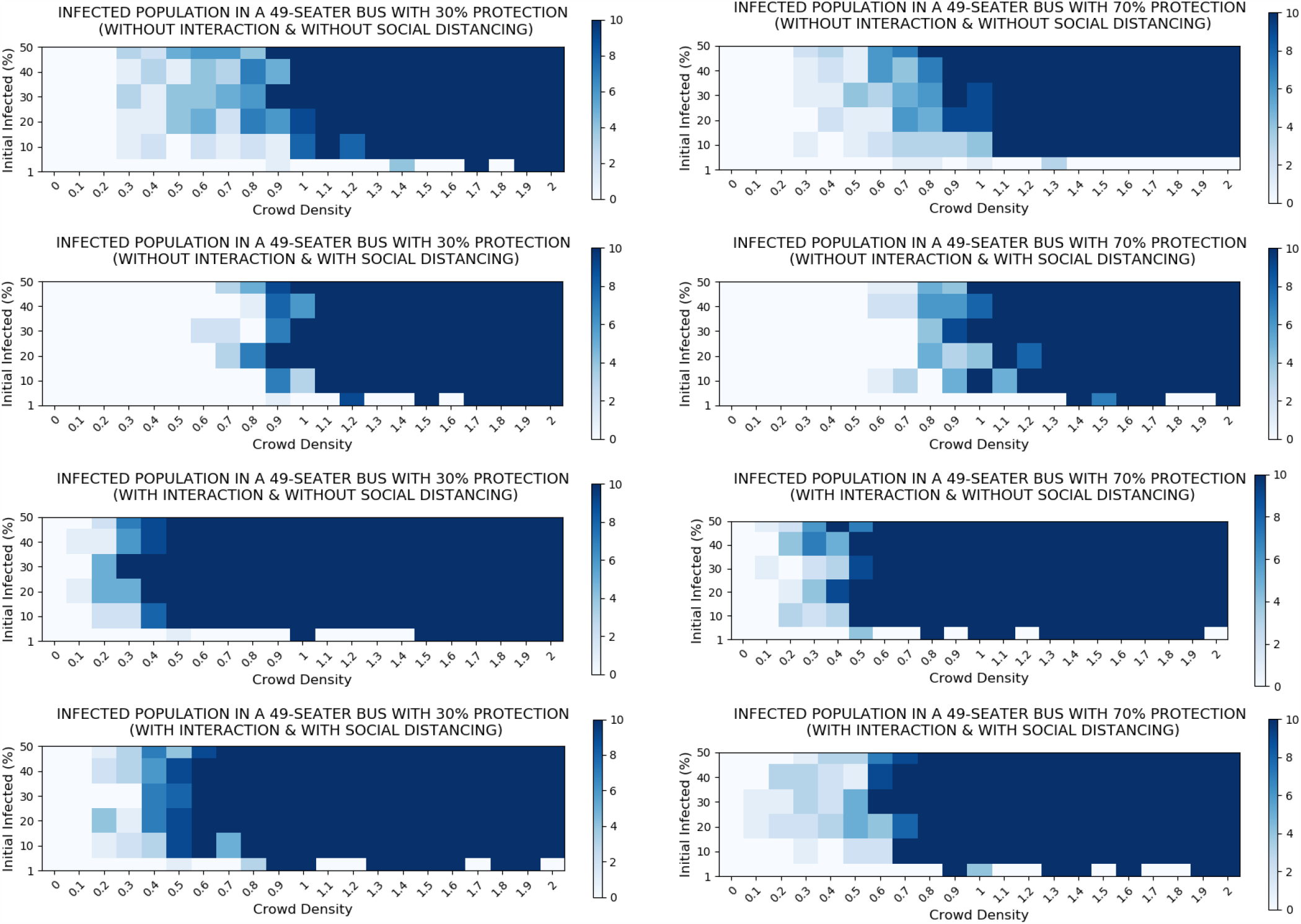
Inside a bus: Resulting final number of newly infected individuals based on crowd density of passengers and the initial percentage of infected individuals (in %), with protection *=* 30% (left column), and *=* 70% (right column).

### 3.3. Disease Transmission on Both Systems

The two transportation systems exhibited identical behaviors when protection level and crowd density were varied. In this section, we look at the number of new infected individuals and the simulation time before reaching the maximum number of infected individuals, given a fixed condition on both systems. We consider the case with 50% protection level and 50% crowd density where both social distancing and interaction are allowed.

#### 3.3.1. Number of Newly Infected Individuals

We look at the final number of new infected individuals in both transportation systems, as shown in Figure 6 per infected individual per protection level. As shown in the graph, the bus transportation system mostly generates more infections than the train. This may be due to the greater capacity of the said transportation system. The greater the occupied capacity, the greater the number of infected individuals since there are a greater the number of susceptible individuals. This is true for all social distancing and interaction scenarios.

**Figure 6:**
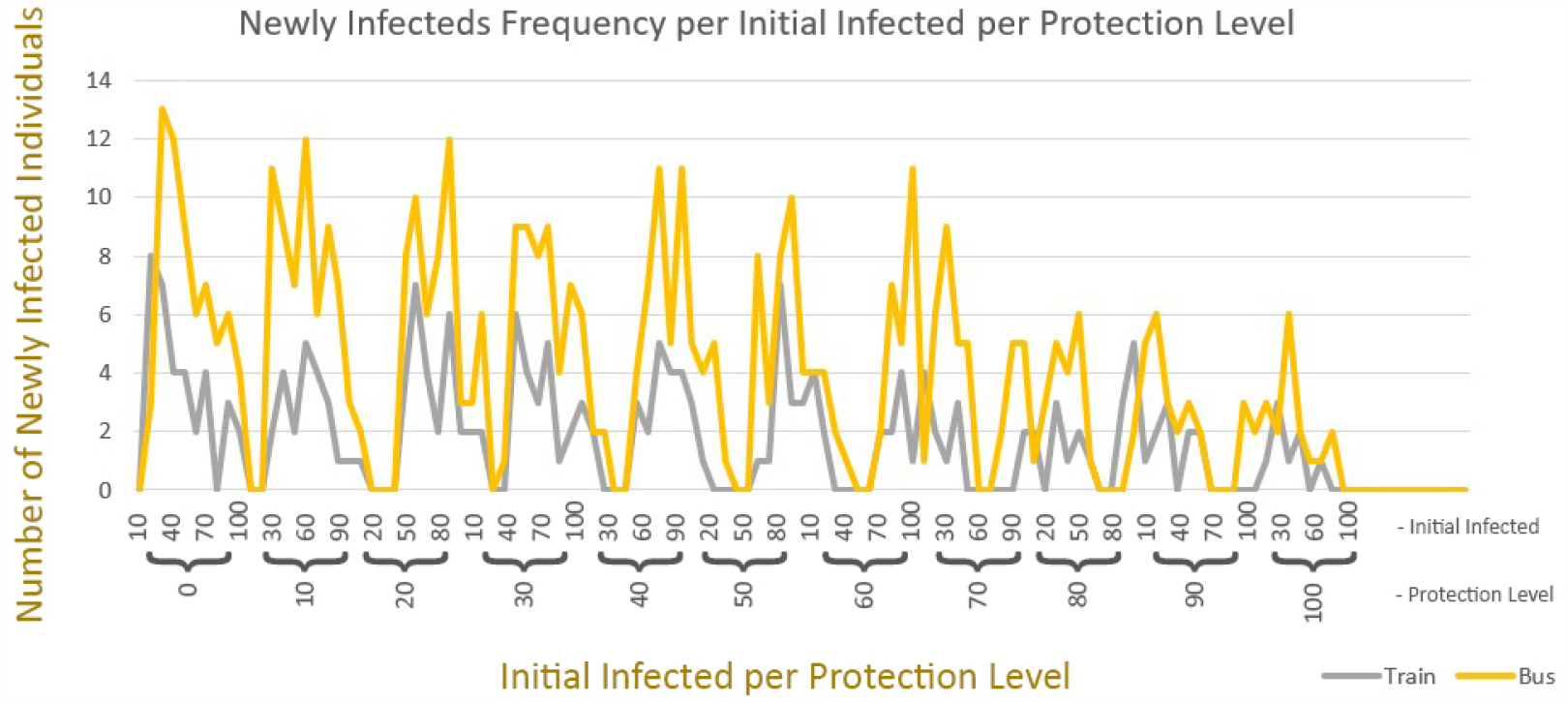
Frequency of new infected individuals for both train and bus scenarios, with protection *=* 50% and crowd density *=* 50%.

**Figure 7:**
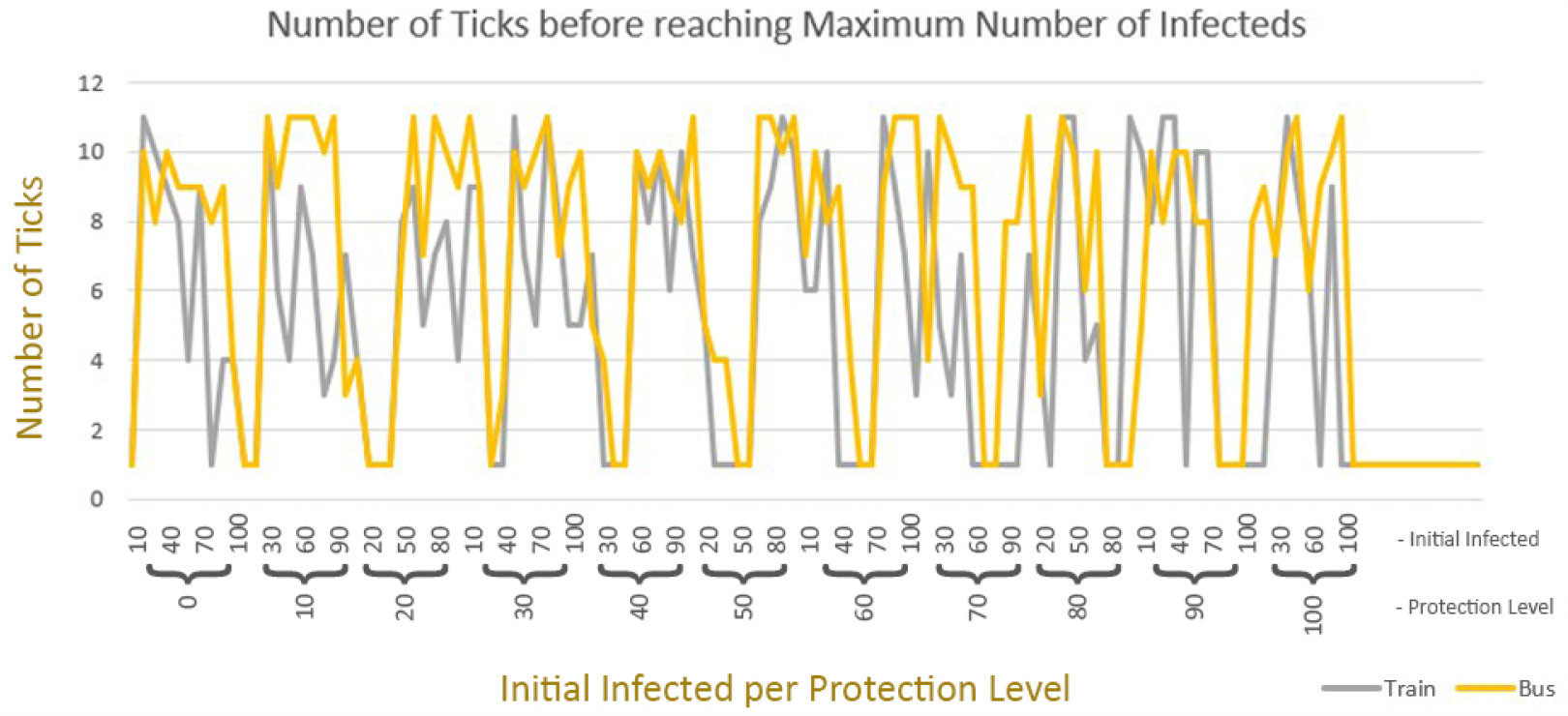
Rate of transmission (through number of ticks) for both the train and the bus scenarios, with protection *=* 50% and crowd density *=* 50%.

#### 3.3.2. Rate of Infection

We assume 1 tick *=* 10 minutes in this simulation. We now observe the number of ticks needed for every setting to reach the final number of infected individuals based on the simulations and assumptions in Figure 6. We observe that in most scenarios the transmission in the train takes shorter time to reach its maximum compared to the bus. This may be due to the smaller size of the train wagon. Thus, the smaller the dimensions of the transportation system, the faster the transmission of the infectious disease. This is true for all social distancing and interaction scenarios.

## 4. Conclusions

In this study, an agent-based model was used to determine the effects of crowd density and protection in the spread of a respiratory infection in mass transportation systems, such as in a 49-seater bus and a 24-seater train wagon. We considered all combinations of scenarios involving the implementation of social distancing and interaction among individuals. We tested different values of parameters that affect the transmission of respiratory infectious disease inside a closed space for all scenarios. These parameters are the protection level of individuals, crowd density inside the mass transportation system, and initial percentage of infected individuals. We then analyzed the effect of these parameters to the number of newly infected individuals.

In terms of protection, results in both the train and bus systems showed that given any percentage of infected individuals, the number of newly infected individuals decreases as the level of protection increases. Having a protection level of 90% or higher is necessary if an individual will use these transportation systems. Moreover, as the crowd density increases, the greater the number of individuals is expected to be infected. In order to prevent spread of infection, crowd density should be less than 10%, but may be increased depending on the preventive measures being applied.

Application of social distancing on our simulations proved to be an effective measure to decrease the number of newly infected individuals. Same goes with limiting the interaction among individuals. Simulations with social distancing or with physical distancing of at least 1 meter and no movement or interaction among passengers showed the least number of infections on both systems, and is the ideal set-up for any of these systems. When these practices are applied, passenger capacity for both the bus and train can be up to 50% while maintaining a minimum number of infections that will occur. This means that a train wagon should have at most 12 passengers while a bus should have at most 24.

We also observed the behavior of transmission between these two systems on a fixed scenario, with fixed parameter values. Given the larger capacity of bus compared to a train wagon, the bus scenario generated more infections during the simulation. However, the infections inside the train was faster in reaching its peak number of infections, due to a smaller dimension. It is easier to encounter an infected individual in a smaller transportation system. A decrease in the crowd density on larger vehicles, or decrease in travel time inside smaller transportation systems may be looked upon by decision makers to address the epidemic.

Overall, our study demonstrated that inside a bus or a train, practice of social or physical distancing and minimizing movement or interaction among passengers are necessary to reduce the risk of being infected. With the preventive measures being practiced, mass transportation systems may take passengers up to 10-50% of its maximum seating capacity. A 10% crowd density with individuals with protection of 90% effectiveness is an ideal set-up to inhibit transmission of respiratory infectious diseases, such as SARS-CoV-2. Further analysis of our models can aid decision makers in designing a policy during times of pandemics or disease outbreaks, in order to protect individuals while maintaining their mode of transportation.

## 5. Limitations

The model considers the dynamics of individuals only when inside the train wagon or bus. This means that, for example, queuing dynamics at the train station or at the bus stop are not included in the model. It should be noted that as the crowd density is decreased inside each transportation system, more trips are needed to ferry passengers, and the duration between both the train schedules and bus schedules should also be minimized to decrease the spread of infectious diseases such as COVID-19 at the station platforms. The inflow and outflow of passengers at various destinations are also not explicitly included in the model.

## Data Availability

Data used in the study are available upon request.

## 6. Supplementary Files

The NetLogo files can be found online at: https://github.com/alvinizer/COVID19NLogoSimulations and sample simulations can be viewed via Youtube:

- Train Simulation: https://youtu.be/AjArjYpTKk
- Bus Simulation: https://youtu.be/syrau2mmHdk

## Acknowledgment

JFR is supported by the Associate Scheme of the Abdus Salam International Centre for Theoretical Physics, Italy. Authors are members of the University of the Philippines COVID-19 Pandemic Response Team.

The authors declare no competing interests.

